# Effects of executive function training on balance and auditory-cognitive dual-task performance in adults with and without hearing loss

**DOI:** 10.1101/2025.08.15.25333755

**Authors:** Niroshica Mohanathas, Lianna Montanari, Rachel Downey, Karen Z. H. Li, M. Kathleen Pichora-Fuller, Louis Bherer, Maxime Lussier, Natalie Phillips, Walter Wittich, Nancy St-Onge, Jean-Pierre Gagne, Jennifer L. Campos

## Abstract

**Background:** Multitasking, such as listening while balancing, relies on integrated processing in the sensory, cognitive, and motor systems; systems that often decline with age. Hearing loss is linked to increased risks of both falls and cognitive decline. Improving cognitive processing through executive function (EF) training may support balance, especially in older adults with hearing loss. This randomized controlled study examined the effects of a 12-week EF training program (targeting inhibition, task switching, divided attention, working memory) on postural outcomes in middle-aged adults with normal hearing (MA; *n* = 19), older adults with normal hearing (OA; *n* = 23), and older adults with hearing loss who used hearing aids (OAHL; *n* = 23), using a dual-task paradigm in a realistic virtual reality environment.

**Methods:** Sixty-five participants were randomly assigned within each age group to an EF training condition or a control condition. Primary outcome measures were auditory-cognitive reaction time on an auditory 2-back working memory task and postural measures (center of pressure path length variability), which were collected in single- and dual-task conditions. Secondary analyses examined whether sensory, cognitive, and mobility performance, as evaluated by baseline standardized assessments, predicted training-related outcomes.

**Results:** Across MA, OA, and OAHL groups, cognitive performance generally improved following EF training and transfer of these training effects were observed during experimental postural tasks and auditory-cognitive tasks, but differed depending on age, hearing thresholds (pure-tone audiometry), and cognitive abilities. Specifically, for postural outcomes, performance improved after training, but only for older adults with better hearing, while those with poorer hearing (at any age) did not improve. For auditory-cognitive task performance, older adults with the poorest hearing and cognition benefited the most from training.

**Conclusions:** EF training may support balance and cognition in older adults, although its benefits for balance may be limited by severe hearing loss, underscoring the value of early intervention.

Trial: https://clinicaltrials.gov/ct2/show/NCT05418998

## Introduction

### Associations among hearing loss, cognition, and mobility

Multitasking in our everyday lives involves successfully integrating and coordinating information from our sensory, cognitive, and motor systems (e.g., when listening, looking, and thinking while moving). Age-related changes across these systems are common [1–6]. In older adults, clinically significant impairments in one domain (e.g., hearing) are also often associated with declines or impairments in the other domains (i.e., cognition and mobility) [7]. For example, hearing loss is associated with cognitive decline [1, 7–14] and dementia [7–8, 10–11, 14] with greater hearing loss severity linked to a greater risk of dementia [10,15]. Hearing loss has also been identified as a top potentially modifiable mid-life risk factor for dementia [10]. Separately, other research has demonstrated that hearing loss is associated with mobility-related problems, including postural instability [16-20, although see 21 for a critical review], slower walking pace [22], elevated vestibular perceptual thresholds [23], poorer physical functioning [24], and a higher likelihood of falls [22, 25–28]. In fact, there is evidence that older adults with hearing loss have a threefold higher risk of falling compared to older adults with normal hearing [28], with a recent study reporting a 17% increased longitudinal risk and 51% increased cross-sectional risk of falls in older adults with hearing loss compared to older adults with normal hearing [29]. These associations among hearing loss, cognitive decline, and mobility problems may reflect interactive mechanisms. Even among older adults with normal hearing and cognitive decline, poorer executive functioning (EF), in particular, is known to be associated with poorer mobility-related outcomes [30–35]. These findings suggest that cognitive mechanisms may also partially underlie the association between hearing loss and poor mobility given that high cognitive load during effortful listening, may a) limit the remaining cognitive resources available to support other concurrently performed tasks (e.g., safe balancing or walking) [26, 36–37], and/or, b) may be associated with changes in brain structure and functioning over time that negatively affect cognitive abilities [37–38]. According to the Framework for Understanding Effortful Listening model, effortful listening may draw on domain-general cognitive resources, particularly under degraded auditory conditions [39] and this could be detrimental to other important tasks that draw on similar resources, such as walking and balancing [40–41].

### Dual-task paradigms to evaluate the effects of the competing demands of hearing and cognition on balance and mobility

Dual-task paradigms are often used to evaluate how cognitive resources are distributed when managing hearing and competing mobility task demands (i.e., gait, balance, walking; see 42 and 43 for reviews). In the context of understanding the interactions and the distribution of cognitive resources among hearing, cognition, and mobility tasks, dual-task paradigms have introduced auditory and/or cognitive tasks that are performed during standing balance or walking tasks. Specifically, single-task auditory-cognitive performance (listening, counting, or remembering) and single-task mobility-related performance (e.g., standing, walking) are compared to dual-task performance (e.g., standing or walking while performing an auditory-cognitive task simultaneously) to determine task prioritization and allocation of cognitive resources [32, 35, 41, 44–45]. For example, in a dual-task study comparing older and younger adults with normal hearing, Carr et al. [46] used a listening-while-balancing task and found that, compared to younger adults, older adults exhibited poorer balance when listening demands were high on a repeating words in noise task [46]. Nieborowska and colleagues [47] implemented a listening-while-walking dual-task experiment and demonstrated worse listening performance (on a multi-talker word identification task) and greater postural prioritization in older compared to younger adults [47]. Further, Li et al. (2001) used a memorizing-while-walking dual-task and showed that older adults experienced more dual-task costs on memory performance compared to walking performance, suggesting prioritization of walking [48]. Our previous study examining dual-task performance in older adults with and without hearing loss [49] implemented a listening-while-balancing task and found higher dual-task costs in conditions of lower task demands for older adults with hearing loss compared to older adults with normal hearing [49]. Bruce et al. [50] implemented a dual-task paradigm whereby younger and older adults with normal hearing and older adults with hearing loss completed a working memory task during standing balance perturbations [50]. Individuals with hearing loss experienced greater dual-task costs (i.e., worse cognitive performance in dual-task compared to single-task conditions in the presence of noise) than younger adults and older adults with normal hearing [50]. Lau et al. [51] used a listening-while walking dual-task to test older adults with and without hearing loss and demonstrated worse performance on a multi-talker word identification task in the single- and dual-task conditions and greater stride time variability in older adults with hearing loss compared to those with normal hearing in the dual-task condition [51]. Taken together, the evidence that older adults, and particularly older adults with hearing loss, experience difficulty managing simultaneously auditory, cognitive, and balance-related tasks, indicates that an important next step is to explore whether targeted interventions could mitigate these challenges and improve their dual-task performance.

### Hearing and cognitive interventions to support mobility

Several strategies have been proposed to improve balance in older adults with hearing loss, with specific interventions addressing different hypothesized mechanisms. One such intervention is hearing aids. Hearing aids may amplify important sound signals from the environment that can support mobility, and/or mitigate cognitive load by reducing listening demand, thereby preserving more cognitive resources that may be allocated to support mobility. However, the literature is mixed with respect to the effectiveness of hearing aids on improving balance and mobility, with some studies demonstrating positive effects [52–57], and other studies showing no or minimal benefits [49, 58–62]. As such, hearing aids may not be an adequate solution for improving mobility and mitigating falls risk. Other multimodal interventions should be considered to support mobility in older adults with hearing loss. An alternative intervention is to more directly address the cognitive load hypothesis through cognitive training to increase the availability of cognitive resources for other tasks. Understanding how the effects of sound amplification from hearing aids can be further augmented through cognitive training to improve balance has not been investigated previously.

Cognitive training refers to standardized activities involving structured practice on tasks targeting various cognitive abilities (e.g., working memory, EF, attention, processing speed) with the aim of improving overall cognitive performance [63–65]. The literature on the benefits of cognitive training interventions is mixed, with strong evidence showing improved performance on the specific tasks that are trained, weaker evidence for the transfer of training to similar tasks, and very limited evidence that training benefits broader cognitive abilities or everyday functioning [66; see 67 and 68 for reviews]. In the context of balance and mobility, training EFs may be more promising due to functional overlap with gait/balance. Specifically, training-related improvements in EF following training, particularly those involving attentional control and inhibitory processing, can lead to improvements in gait and balance performance in older adults [32], including single-support balance, sit-to-stand, dynamic posturography, and a 40-foot walk test [69]. Downey et al. [70] and Pothier et al. [71] used the EF training program developed by Lussier et al. [72] that specifically targets different aspects of EF (i.e., inhibition, task switching, divided attention, and working memory) and have shown improvements in cognitive and mobility task performance, including more accurate 2-back-while-walking performance [70] and faster spontaneous walking speed [71]. It has been proposed that EF training may improve the ability to engage in compensatory scaffolding (the recruitment of additional neural resources to compensate for age-related structural and functional brain decline) [32, 73] such that the likelihood of transfer to untrained tasks increases when there is shared neural circuitry between the trained functions and the targeted outcomes [74]. Using EF training to increase the cognitive resources available for multitasking [75] could support better management of simultaneous task demands in older adults with and without hearing loss.

For older adults with hearing loss, increased availability of cognitive resources gained from EF training could reduce competition amongst cognitive and neural resources to better manage greater listening demands associated with hearing loss. Very few studies, however, have evaluated the benefits of EF training on balance and mobility-related outcomes in older adults with hearing loss specifically. Bruce et al. [76] implemented exercise (i.e., cycling) and cognitive training (i.e., computerized, visual discrimination dual-task) interventions simultaneously or sequentially in older adults with and without hearing loss and found that both groups improved in single- and dual-task cognitive performance (especially during low task demands) and sit-to-stand performance under dual-task conditions, but without improvements in balance (i.e., on measures from computerized dynamic posturography including double support, single-support, and visual sway referenced). However, given the need to carefully allocate cognitive resources as a function of multiple demands, it is likely that any effects of EF training would be most pronounced during balance and mobility-related tasks that are complex and multisensory (involving looking, listening, remembering, and moving at the same time). Further, these types of complex scenarios are reflective of everyday life, and it is important to understand whether EF training-related benefits generalize to these types of ecologically valid contexts [77–78]. Previous studies have strategically introduced these challenges using virtual reality (VR) and simulation technologies, which enable the study of complex, realistic situations under safe and controlled conditions [46–47, 49, 51]. However, no previous VR-based studies have evaluated the effects of EF training on balance-related outcomes in older adults with hearing loss.

When implementing an EF training program as a strategy to mitigate age-related declines in cognitive processing, an additional important consideration is the point at which it is the optimal time to intervene, recognizing that training-related effectiveness is likely influenced by age, stage of cognitive changes/decline, and hearing loss trajectory. In fact, declines in domains of cognitive functions such as working memory and speed of processing can begin as early as mid-life [79–83]. Declines in hearing can also begin in mid-life, with nearly 20% of middle-aged adults (45-65 years old) having elevated high-frequency hearing thresholds, and 5% of middle-aged adults having mild hearing loss [84–86]. While very few studies have specifically investigated the associations between hearing and mobility-related outcomes in middle-aged adults, a recent study by Wang et al. [87] found that poorer hearing abilities (defined using audiometric pure-tone average (PTA)) were associated with worse postural control in adults aged 40-69 years. Importantly, even with clinically normal audiometric hearing thresholds, supra-threshold declines (difficulty interpreting auditory information) [88] in hearing (e.g., difficulties understanding speech in noise) often starts in middle age [89–90]. These supra-threshold declines could draw cognitive resources in more subtle ways, as it may result in more effortful listening, thereby introducing greater processing demands (compared to when auditory processing is normal) [90]. Therefore, comparing the effects of cognitive training on mobility-related outcomes in middle-aged adults could, provide unique insights into the effects of EF interventions across the adult lifespan.

### Current research

EF training has the potential to improve cognitive processing, thereby improving balance-related outcomes. The most pronounced balance-related effects of EF training are likely to be observed during complex, ecologically valid contexts that mirror the demands of everyday life. These training-related benefits may start in middle age, increase with older age, and may be more pronounced in older adults with hearing loss, who may be at risk for declines in EF, mobility problems and falls. It is also unknown whether EF training can further augment any benefits already provided through amplification. Therefore, this randomized control study evaluated the effects of a 12-week EF training program on middle-aged and older adults with normal hearing (MA and OA, respectively) and older adults with hearing loss who use hearing aids (OAHL). Pre- and post-training effects (T1 vs. T2) were evaluated using a dual-task paradigm (auditory-cognitive + balancing tasks) in an immersive, realistic, VR environment. Secondary analyses also examined whether sensory, cognitive, and mobility functions, as evaluated by baseline standardized assessments, predicted training-related outcomes.

## Methods

### Participants

Nineteen MA (*M_age_* = 52, *SD_age_* = 5; 11 EF training, 8 controls; 14 F, 5 M), 23 OA (*M_age_* = 70.87, *SD_age_* = 3.83; 13 EF training, 10 controls; 13 F, 10 M), and 23 OAHL (*M_age_* = 76.48, *SD_age_* = 6.48; 14 EF training, 9 controls; 9 F, 14 M) participated in this study (see Fig 1 for consort diagram). Recruitment strategies included both advertisements posted across Toronto and digital outreach via social media. Participants were included in the MA group if they were 45-60 years old and had normal audiometric hearing thresholds, the OA group if they were 65+ years old and had normal audiometric hearing thresholds, and the OAHL group if they were 65+ years old, had bilateral hearing loss and had used hearing aids for a minimum of 6 months. While this ensured that participants were comfortable with and well-adapted to their hearing aids, there was variability in the length of time participants used their current hearing aids (*M*= 2.14 years, range = 1 week - 5 years), frequency of use (*M* = 13.32 h/day, *SD* = 5.10, range = 3–24), fit (hearing had been assessed by an audiologist within the previous 2 years on average), and model type (*n* =14 behind-the-ear; *n* = 3 in-the-canal; *n* = 2 in-the-ear). Hearing thresholds were measured using SHOEBOX^TM^ [91]. A 4-frequency PTA was calculated for each ear using thresholds at 0.5, 1, 2, and 4 kHz. Participants with a better-ear PTA of ≤ 25 dB HL were classified as having normal hearing and those with a PTA ≥ 26 dB HL were classified as having hearing loss [92]. While the updated World Health Organization [93] standard cut-off is <20 dB HL and has been used in current studies [e.g., 94], we used the previously recommended cut-off of ≤ 25dB HL [93] to be comparable and consistent with previous research in this field. All participants had symmetrical hearing, defined as a right-left ear threshold difference of no more than 15 dB at two or more adjacent frequencies. Participants also had to speak and have learned English fluently before the age of five and have normal or corrected-to-normal visual acuity with their habitual correction on the Early Treatment Diabetic Retinopathy Study chart (ETDRS; logMAR of -0.2-0.5 in the better eye) [95–96]. All participants were required to have normal cognition as determined through screening using the Montreal Cognitive Assessment (MoCA) [97], with a ≥23 cutoff off [98]. See S1 for detailed descriptions of all standardized assessment measures. Participants were excluded if they had a self-reported history of neurological or psychiatric illness, mild cognitive impairment or dementia, recent stroke, seizure disorder or epilepsy, claustrophobia, susceptibility to motion sickness, arthritis, or a serious injury affecting the hand or arm (as the responses during the experiment were made using a game controller). Additional exclusion criteria included mobility-related impairments (e.g., use of mobility aids, diagnosed vestibular disorders, or major musculoskeletal conditions that interfere with daily activities) and current participation in any other cognitive training or exercise programs. This study was approved by the University Health Network’s research ethics board (Study ID: 19-5857). Data collection began June 12th, 2023, and was completed by April 2nd, 2024. Participants were compensated a minimum of $220 ($30 for the four in-person sessions and $100 for the 12-week at-home training period). See published protocol [99] and registered clinical trial (https://clinicaltrials.gov/ct2/show/NCT05418998).

**Fig 1.**
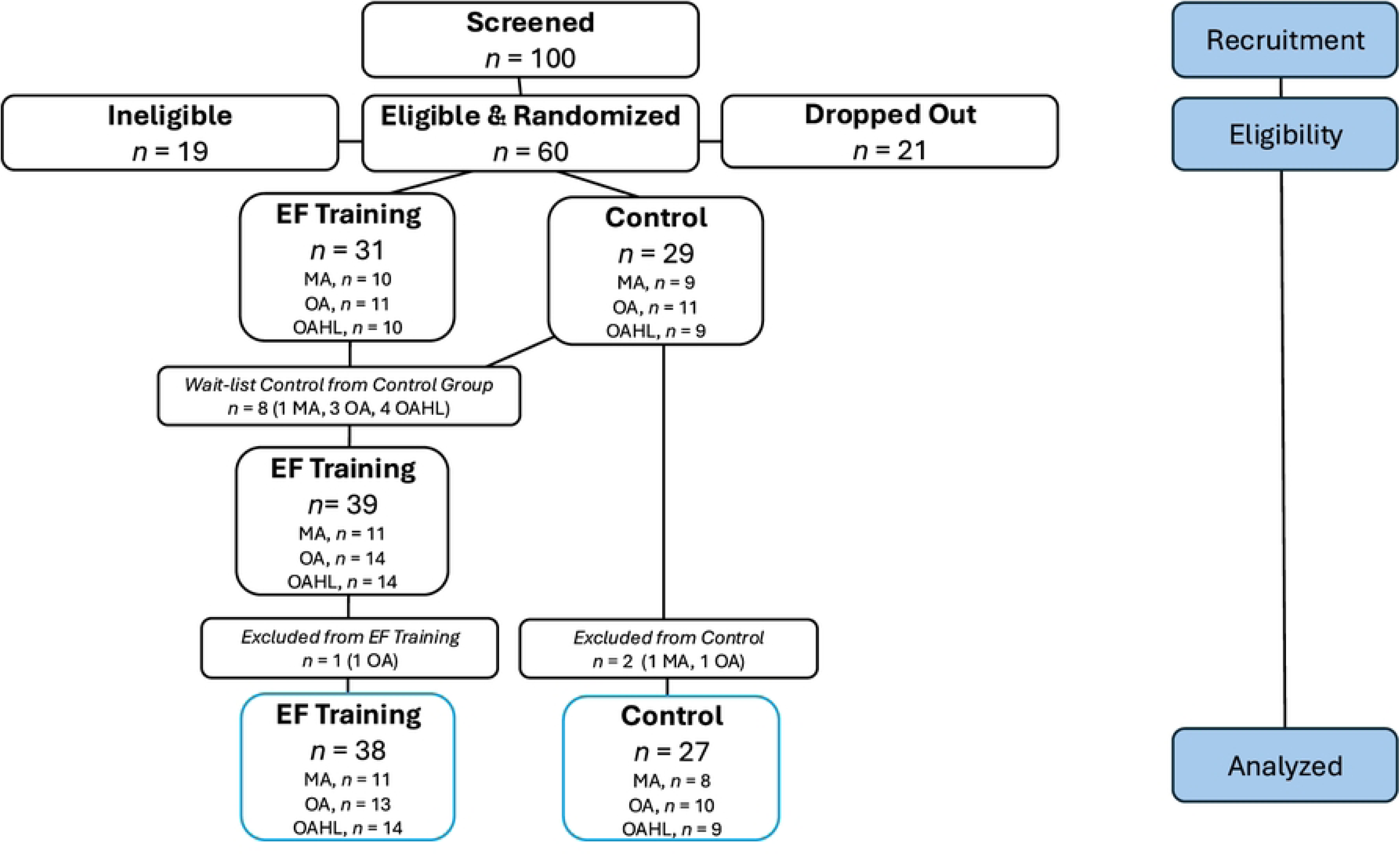
Consort Flowchart of Participant Recruitment, Eligibility, and Assignment.

**Fig 1**. Consort diagram of participant recruitment and the final eligible sample analyzed after exclusions and dropouts. Specifically, one hundred participants were screened, sixty of whom were eligible. Nineteen participants were ineligible, and twenty-one individuals dropped out of the study due to various reasons (i.e., sickness including COVID-19, could not commit to study time commitment, no longer interested in participating, travel abroad). Of the sixty eligible participants, thirty-one were randomly assigned to the executive function (EF) condition, while twenty-nine were assigned to the control condition. Eight participants who were originally assigned to the control condition came back to complete the 12-week training and were added to the EF training condition, bringing this sample size of the EF training group to thirty-nine. Three participants (one from the EF condition and two from the control condition) were excluded due to technical errors that resulted in missing data. The final sample analyzed included thirty-eight participants in EF training and twenty-seven controls across the three age groups (blue outlined boxes on the last line). MA = Middle-Aged Adults with normal hearing, OA = Older Adults with normal hearing, OAHL = Older Adults with Hearing Loss.

### Study design and procedure

First, a screening was conducted by telephone to determine a participant’s initial eligibility to participate in the study. The screening included demographic and self-reported responses regarding eligibility criteria, as well as questions about the participant’s level of comfort in using a computer or tablet at home to complete the computerized training. Eligible participants were then asked to come to KITE-Toronto Rehabilitation Institute, University Health Network to provide written consent and complete a battery of sensory, mobility, and cognitive assessments (secondary measures of interest at T1 and T2, see below). If participants met the screening criteria (the pre-determined cut-offs for the ETDRS and MoCA measures, and the age and hearing criteria for inclusion in one of the MA, OA, or OAHL groups), they were invited to complete the experimental session.

The experimental session involved completing an auditory-cognitive task (i.e., 2-back task) and a standing balance task (each task alone and simultaneously) in an immersive VR simulator (see procedural details below). Participants were then randomized into either the EF training condition or the control condition for 12 weeks, with an attempt to balance the number of males and females within each group. After 12 weeks, participants returned to the lab and completed the same set of secondary measures (see Downey et al. [99] and below for details) and experimental tasks again at T2. Upon completing the study (T1 and T2), participants in the control condition were then offered access to the at-home EF training. A subset of the participants in the control condition (*n* = 8; see Fig 1) chose to complete the 12-week EF training intervention and following training they were brought back to the lab to complete the full post-training protocol. This waitlist-control subset included one MA, three OA, and four OAHL participants. These eight participants did not show any differences in performance compared to the participants in the original training condition and therefore their initial data were included in analyses for the control condition and their post EF training data were included in the analyses for the training condition (see Fig 1).

### Characterizing the participant sample and secondary outcome measures

To address the objectives of determining whether sensory, cognitive, and mobility-related abilities predict training-related outcomes and to further characterize the participant sample beyond eligibility criteria (aside from the PTA, MoCA and ETDRS measures), additional assessments were conducted in-person or online via Qualtrics (an online survey software; Qualtrics, Provo, Utah).

#### Hearing

The Canadian Digit Triplet Test [100], Hearing Handicap Inventory for the Elderly [101] and Listening Self-Efficacy Questionnaire [102] were administered.

#### Cognition

The Rey Auditory Verbal Learning Test [103], Digit Span Forward & Backward [104], Letter Number Sequencing [104], Digit Symbol-Coding [104], Stroop Color-Word Interference [105], Trail Making Test [106] and Frequency of Forgetting Questionnaire [107] were administered. ***Vision.*** The Pelli-Robson Contrast Sensitivity Test were administered [108].

#### Mobility

Mini-BESTest [109] and Activities-Specific Balance Confidence Scale [110] were administered.

#### Other

The Health History Questionnaire (see Table 1 for a summary of demographics and S1 for assessment descriptions) were administered. All assessments were administered again at T2 except for PTA, Hearing Handicap Inventory for the Elderly, ETDRS and Health History Questionnaire.

**Table 1.** Demographics Table of Baseline Sensory, Cognitive, and Mobility Function.

Table 1. All sensory, cognitive and mobility assessments completed at T1 in-person or online via Qualtrics, separated by Group, Middle-Aged Adults with normal hearing (MA), Older Adults with normal hearing (OA), and Older Adults with Hearing Loss (OAHL). One-way ANOVAs were conducted for each measure to determine if there were any differences across groups and *p*-values for main effects, with significant post-hoc comparisons listed (bolded = significant). ^1^ Canadian Digit Triplet Test, Speech Reception Threshold in decibel signal-to-noise ratio (SRT in dB SNR). ^2^ Early Treatment Diabetic Retinopathy Study Acuity logarithm of the Minimum Angle of Resolution (logMAR). ^3^ Binocular Pelli Robson Contrast Sensitivity logarithmic Contrast Sensitivity (logCS).

### Experimental session and primary outcome measures

#### StreetLab VR simulator

The experimental session was completed in StreetLab, a fully immersive, state-of-the-art, multisensory, projection-based, VR simulator used to simulate realistic conditions (see Fig 2). StreetLab has a 240° horizontal by +15° to -90° vertical field-of-view curved projection screen extending from floor to ceiling. The virtual environment used in this study depicted a large, urban, 6-lane intersection street-crossing in Toronto (developed using OpenScene Graph www.openscenegraph.org). StreetLab is outfitted with an AMTI (Advanced Mechanical Technology, Inc., Watertown, MA) BP12001200–2000 strain gage force plate that measures ground reaction forces. A harness was used to ensure participants could not fall to the ground surface but did not provide body-weight assistance during the study while standing. There is a spatialized surround sound system with seven speakers (Meyersound MP-4XP; Meyersound Laboratories, Inc., Berkeley, CA) distributed behind the projection screen, at approximately the height of a participant’s head when they are standing on the force plate positioned at 0**°** azimuth across a horizontal plane at ± 28° (right front and left front), ± 90° (right side and left side), and ± 127° (right rear and left rear). One subwoofer (Meyersound MP-10XP) is located in the floor under the centre speaker. All loudspeakers are at a distance of 2.14 m from the participant when standing on the force plate. A sound level meter recorded ambient noise in the laboratory at 51 dB SPL during quiet standing conditions (full acoustic details of the StreetLab environment are available in Campos et al. [111]).

**Fig 2.**
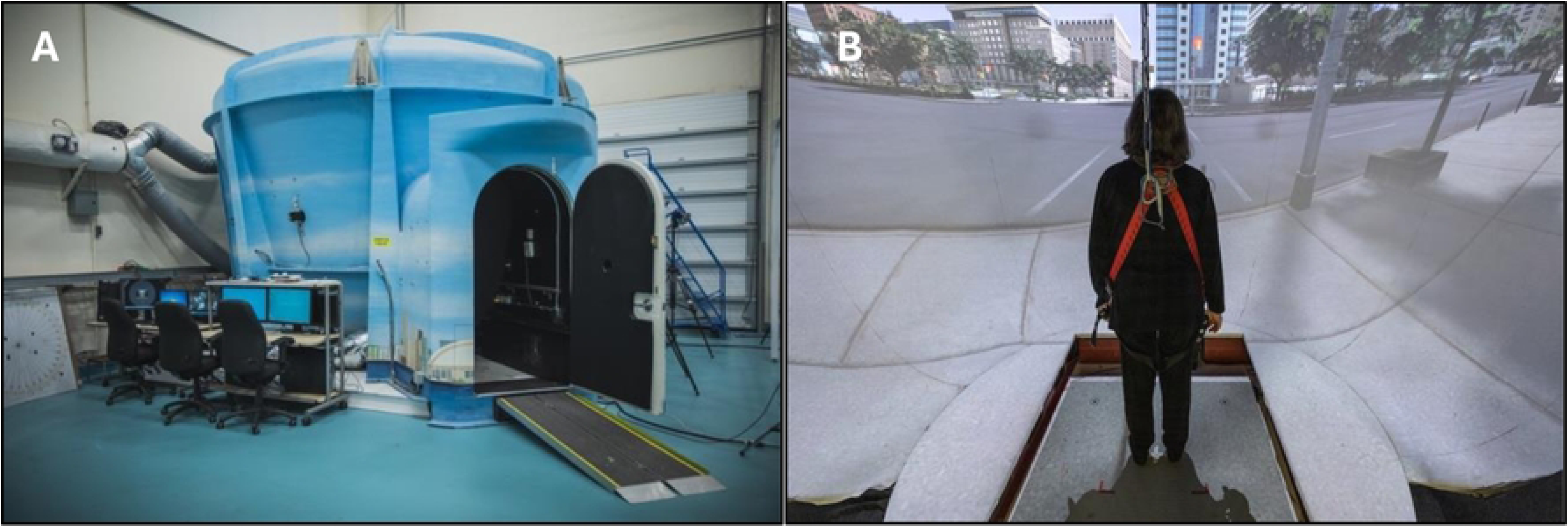
StreetLab.

**Fig 2**. (a) Exterior view of StreetLab. (b) Interior view of StreetLab depicting a participant standing on the force plate, oriented toward the projection screen displaying a simulated city street-crossing.

#### Auditory 2-back working memory task

An auditorily presented, 2-back version of the *n*-back working memory task was implemented during single- and dual-task conditions [112]. Before the experiment began, the amplitude of the stimuli was set to 60 dB A (similar to the level of typical conversational speech) during an auditory 1-back task. For the 1-back task, participants were presented with 15 single digits sequentially and asked to repeat each digit as soon as it was presented. To confirm all participants could perform the task, they were required to achieve an accuracy of 100% on the task, which was achieved by all participants. Therefore, the stimuli were set to 60 dB A for the remainder of the experimental session. Participants next completed three to four practice lists of 15 single digits in an auditory 2-back task, where they were instructed to indicate whether the number they were presented with was the same or different than the number that was presented 2 steps (numbers) previously. They responded “same” using the right button or “different” using the left button on a handheld gaming controller (Forty4 Wireless Gaming Controller) and were required to achieve a minimum accuracy of 70% for at least one of the three practice lists for the 2-back task. For the main experimental task, participants again completed 15 single digits in the 2-back task per condition (see condition descriptions below). The primary auditory 2-back outcome measure was a reaction time weighted (RTW) score. Specifically, RTW (ms) measured the average reaction time across trials, wherein for correct trials, the reaction time was unadjusted, but for incorrect trials, a maximum of 2000 ms was given (the maximum duration for each trial) to use a metric that accounts for both RT and accuracy [113–114].

#### Standing balance task

During single- and dual-task standing balance conditions, participants stood with their feet shoulder-width apart on the force platform with their arms at their sides for approximately 60 seconds [115], of which the first 48 seconds were used for data analysis to ensure equal trial durations across all participants and conditions. The primary postural outcome measure was Centre of Pressure (COP) path length variability (standard deviation of path length, mm) in the anterior–posterior direction (APSD, front and back sway). Participants completed the standing balance task with their eyes open (easier) and eyes closed (harder) to manipulate postural difficulty. To control for variations in hand positioning that could affect postural outcomes between single-task standing and dual-task conditions, participants were instructed to hold the gaming controller during the single-task standing condition even though no button responses were required. Participants completed the three conditions in an ABC-CBA sequence (A: single-task standing, B: single-task auditory 2-back, and C: dual-task). This occurred twice, once with eyes open and once with eyes closed, counterbalanced for order across participants.

#### Twelve-week EF cognitive training program

Following the completion of the baseline secondary measures, participants were randomly assigned to either the EF training condition or the control condition and were made aware of which treatment condition they were assigned to. This was a single-blind study, such that researchers conducting the pre-training/T1 and post-training/T2 assessments were blind to the participants’ treatment condition assignment. The researchers calling participants during the 12-week program to track their weekly progress (see below) were not the same researchers who conducted the in-lab assessments. Participants assigned to the EF training condition received an online orientation session via Zoom to familiarize them with the EF training program. The participants in the EF training condition then engaged in 12 weeks of at-home training, while participants in the control condition continued their usual activities. To control for potential confounding factors between conditions related to differences in engagement with the study team, both groups received weekly phone calls to track any changes in lifestyle, stress, energy levels, or significant life events. Additionally, during these weekly calls, participants in the EF training condition were asked about their training experience, including any difficulties encountered, general perceived improvements, technical issues, or missed sessions.

The training protocol involved a customized program that included three separate tasks; visual *n*-back, Stroop, and a visual discrimination dual-task (see below for details on each task) [72]. The training program was designed to target multiple components of EF, including inhibition, task switching, divided attention, and working memory. Participants completed 6 training sessions across 3 days per week (two modules, 15 minutes each). Participants were instructed to space their training sessions evenly throughout the week, with approximately 48 hours between each session. They were instructed to take a break between modules but not during a module. The first session during Week 1 and last session during Week 12 were considered the pre- and post-training “evaluation modules” respectively, while the remaining 10 sessions were considered “training modules” (described in detail below). The evaluation and training modules were designed to assess the same domains of cognitive functioning as those being trained; however, the stimuli for the specific tasks differed in order to establish near-transfer effects from training. The computerized training could be completed on a laptop, desktop, or tablet, and each participant used the same device throughout the 12-week period. During training, participants were asked to perform each task as quickly and accurately as possible, with RT (ms) recorded for all three tasks. At the end of each session, participants received visual feedback in the form of a performance graph to encourage engagement and allow for self-monitoring of progress.

### EF training program modules

#### Visual *n*-back

The visual *n*-back module was designed to improve the updating and maintenance processes of working memory. During evaluation modules, participants indicated whether the most recently presented number (ranging from 1-9) matched the number presented one (1-back), two (2-back), or three (3-back) trials earlier. The training modules included letters rather than numbers. As training progressed, the proportion of more difficult 2-back and 3-back trials increased to gradually increase task demands. Notably, the modality of the *n*-back task used during EF training (visual *n*-back) differed from that employed in the experiment (auditory *n*-back), allowing for assessment of cross-modal transfer. RTs were calculated for each task, excluding incorrect trials or RTs exceeding 4 seconds. Average RT were calculated for correct trials of the 2-back condition [72, 99, 116].

### Stroop

The Stroop module was designed to improve inhibitory control and task-switching abilities through five distinct tasks: familiarization, reading, counting, inhibition, and switching. In the evaluation modules during the familiarization task, a single number was displayed on the screen (e.g., “5”) and participants were asked to simply identify the number. In the reading task, the same digit was displayed multiple times in a group, with the digit presented corresponding to the number of digits shown (e.g., 5 copies of the digit “5”) and participants were asked to identify the digit (“5”). In the counting condition, 1-6 asterisks appeared on the screen and participants had to simply count and state the number of asterisks presented. In the inhibition task, participants were presented with a digit (e.g., “3”) that did not match the quantity of that digit (e.g., 5 copies of the number 3) and participants had to indicate the digit that was presented (i.e., 3), rather the quantity of digits presented (i.e., 5). In the switching task, participants were presented with a group of digits (e.g., the digit “5”, 2 times) with a white frame surrounding it on some trials. When the white frame was present, the participant was instructed to report the digit presented (in this case “5”). When the white frame was not present, the participant was instructed to report the quantity of digits (e.g., in this case a “2”). In the training modules, letters formed by asterisks or larger letters formed by smaller letters were used as the stimuli. RTs were calculated for each task, excluding incorrect trials or RTs exceeding 4 seconds. Average RT values were calculated for correct trials of the switching task [72, 99, 116].

### Visual Discrimination Dual-Task

The visual discrimination dual-task module was designed to improve divided attention by requiring participants to engage in a visual discrimination task under varying task loads. In the evaluation modules during single-task trials, participants were told which letter keys on the keyboard or tablet corresponded to a single category of image (e.g., either an animal or space image) and then they were asked to press the corresponding letter key. During dual-task trials, animal and space images appeared on the screen, sometimes at the same time or at other times individually; participants were asked to press the key that corresponded to the presented image, while giving equal importance to both image types (i.e., not favouring the animals or celestial bodies such as a picture of a moon). Blocks included single-only trials, dual-only trials, or a mix of both. As well, participants were asked to avoid responding to both image types at the same time as a strategy when both images were presented. In the training modules, fruits and modes of transportation were used as the stimuli rather than animal and celestial body images. RTs were calculated for each task, excluding incorrect trials or RTs exceeding 4 seconds. Dual-task cost RT was calculated which consisted of a ratio based on the average of both hands across dual- and single-mixed trials, (dual-mixed trial RTs – single-mixed trial RTs / single-mixed trial RTs) [72, 99, 116].

### Post-training/T2 session

Following the 12-week period, all participants again completed the three experimental tasks in StreetLab (single-task auditory-cognitive task, single-task standing balance task, dual-task).

#### Statistical analyses

All analyses were completed using R-studio (Version 2024.12.0+467). Histograms were used to visualize the distribution of all dependent measures (i.e., RTW, COP APSD) and boxplots were used to visualize outliers. If data points were above or below three SDs from the mean, instead of excluding them altogether, winsorization was used to replace these data points with the next most non-extreme value. Outcome measures were normally distributed or near-normally distributed (skewness of -.31 for RTW and .48 for COP APSD) and platykurtic to slightly platykurtic (kurtosis of 2.27 for RTW and 2.26 for COP APSD). However, these skewness and kurtosis ranges were deemed to be within acceptable ranges. To preserve the original structure and interpretability of the dataset, no transformations were applied. Continuous variables were group-mean centered to facilitate interpretation of main effects and interactions.

Welch’s two-tailed, between-subjects t-tests were conducted to confirm that the participants assigned to the EF training condition were not statistically different from those assigned to the control condition at T1 with respect to primary outcomes and secondary measures of sensory, cognitive, and motor functioning. Visual inspection of individual participant data and mixed factorial ANOVAs were conducted to confirm whether the EF training condition improved over the course of the 12 weeks in the evaluation modules for Week 1 versus the evaluation modules at Week 12 for the visual *n*-back, Stroop, and visual discrimination dual-task.

Two linear mixed-effect models (LMM) were conducted for each primary outcome measure: auditory 2-back RTW (ms) and COP APSD (mm). Following the approach outlined by [117], a backward elimination strategy was used to derive the most simplified model. Within-subject differences across cohorts were modeled using nested random intercepts. Fixed effects included the following and their interactions: Group (MA, OA, OAHL), Intervention (EF training, control), Time (pre-training/T1, post-training/T2), Task (single-task, dual-task), Eyes, (eyes-open, eyes-closed), PTA, MoCA, Mini-BESTest and covariates such as Age (continuous variable), Education (years) and Sex (male, female). For continuous significant fixed effects (i.e., PTA, MoCA, Age) there were a range of abilities. For example, for PTA participants across the spectrum of hearing abilities were recruited (normal hearing to hearing loss). Therefore, when reporting significant results of PTA below, the different levels of hearing loss were binned into tertiles across the entire sample (“lower PTAs” were 9 dB HL or lower; “average PTAs” ranged from 10 dB HL to 45 dB HL (average = 28 dB HL); and “higher PTAs” were 46 dB HL or higher; the total range of PTAs in the entire sample was 3 dB HL to 71 dB HL). When reporting significant results of MoCA below, the different levels of MoCA were binned into tertiles across the entire sample (“lower MoCA scores” ranged from 23 to 25 out of 30; “average MoCA scores” ranged from 26 to 28 out of 30 (average = 27); “higher MoCA scores” were 29 or 30 out of 30; the range of MoCA scores across the total sample was 23 to 30 out of 30). When reporting significant results of Age, different Age levels were binned into tertiles across the entire sample (“lower Ages” were 56 years or younger; “average Ages” were 57 to 78 years (average = 67); “higher Ages” were 79 years or older; the range of ages across the entire sample was 45 to 90 years). Post-hoc tests were conducted with Bonferroni correction. LMMs were refined based on likelihood ratio tests evaluating model fit. Non-contributory fixed effects and interactions were excluded unless they represented key confounding variables (e.g., age, education, sex) or were integral to testing the primary or secondary objectives. Note that only significant main and interaction factors of Time or Time x Intervention are reported below, as these significant results allow us to directly evaluate our primary and secondary research questions of interest, which are focused on training-related effects. The threshold for significance was set to *p* < 0.05.

## Results

### Comparability of participants in the control and training conditions

To ensure participants in the EF training and control conditions had similar sensory, cognitive, and mobility abilities at baseline, we conducted t-tests across all three groups between the participants assigned to EF training condition and those assigned to the control condition across all standardized measures and found no significant differences with two exceptions. Specifically, there was a significant difference in age between those in the OA group who were assigned to the EF training condition compared to those assigned to the control condition (*p* = 0.024), with those in EF training condition being younger (*M* = 69.54) than those in the control condition (*M* = 72.60) and Mini-BESTest performance (*p* = 0.041) of those in the EF training condition being significantly worse (*M* = 23.08) than those in the control condition (*M* = 25.10; see Table 1 for demographics across all three groups). To ensure participants assigned to the EF training and control conditions had similar primary outcomes at baseline, we conducted t-tests across all three groups between the participants assigned to the EF training condition and those assigned to the control condition across RTW and APSD and found no significant differences.

### Demonstrated effects of the 12-week training program

To determine whether participants in the EF training condition demonstrated improvements from Week 1 of training to Week 12 of training, the evaluation modules for the visual *n*-back, Stroop, and visual discrimination dual-task RTs were compared. Overall, visual inspection of the data revealed that over the 12 weeks, every individual improved on the visual *n*-back task (apart from 1 OAHL) and the Stroop task (apart from 1 MA). However, the results for the visual discrimination dual-task were mixed, with some participants improving (3 MA, 4 OA, 5 OAHL) and others not improving (7 MA, 9 OA and 9 OAHL). To evaluate these training-related effects statistically, separate mixed-factorial ANOVAs, including the within-subject factor Time (Week 1, Week 12) and between-subjects factor Group (MA, OA, OAHL) were conducted for each of the three training tasks. For the Stroop task and visual *n*-back task, a main effect of Time was observed indicating that Week 12 RT performance was better (i.e., faster) than Week 1 RT performance; (*F*(1, 34) = 47.29, *p* < 0.001 and *F*(1, 32) = 61.82, *p* < 0.001, respectively). However, for the visual discrimination dual-task there were no significant improvements. While a main effect of Time was also observed for the visual discrimination dual-task (*F*(1, 34) = 7.23, *p* = 0.011), the effects were in the opposite direction, such that RT performance was worse at Week 12 compared to Week 1. There were no main effects of Group or interaction effects for any of the three training tasks.

### Effects of EF training on balance and auditory-cognitive performance in StreetLab

#### Standing Balance Performance

To address our primary objective of investigating the effects of EF training across Time and Groups on standing balance performance, an LMM was conducted with the primary balance outcome of interest, COP APSD (mm), as the dependent variable, where higher values were interpreted in this context as indicating worse performance. First, a significant four-way Intervention*Time*Age*PTA interaction was demonstrated on COP APSD performance, *F*(1,425.11) = 5.0022, *p* = 0.026, *η_p_ ^2^* = .01, with post-hoc tests demonstrating that for those in the EF training conditions (but not controls), T2 balance was significantly better (smaller COP APSD) than T1 balance for participants of older ages and lower dB HL levels (i.e., better hearing; *t* = 2.78, *p* = 0.006, Fig 3). Further, for those in the EF training condition, there were no improvements in balance at T2 versus T1 in those with the average to higher levels of hearing loss across all age tertiles (younger, average, older; performance was worse; see S2 for full statistics). To summarize, those in the EF training condition who were of older ages and had lower levels of hearing loss (i.e., better hearing) demonstrated significant improvements in balance between T1 and T2; however, those in the EF training condition with average to higher levels of hearing loss (across all ages) showed no improvements after training.

**Fig 3.**
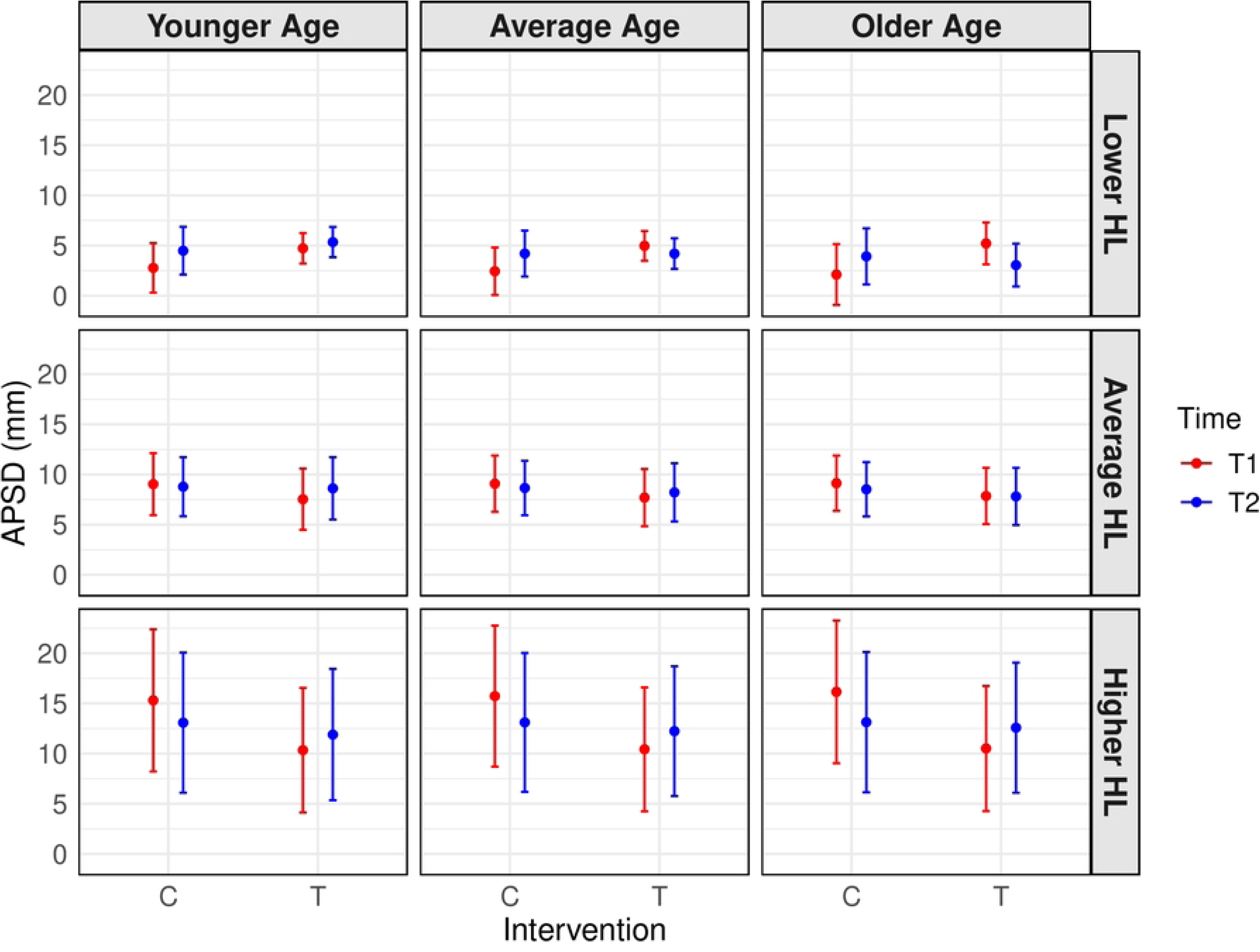
LMM of Intervention x Time x Age x PTA Interaction Effect on COP APSD.

**Fig 3**. Estimated marginal means of Centre of Pressure Path Length Anterior-Posterior Standard Deviation (COP APSD in mm; higher scores represent worse performance) from T1 to T2 for those in both the EF cognitive training (T) and control (C) conditions. Age and hearing loss (HL) tertiles are represented by the columns and rows respectively. All significant findings are stated in-text. Errors bars represent the 95% confidence intervals.

Second, we found a significant four-way Intervention*Time*Group (MA, OA, OAHL)*MoCA interaction on COP APSD performance, *F*(2,426.06) = 8.8496, *p* < .001, *η_p_ ^2^* = .04, but for which no post-hoc tests indicated improvements in balance related to training. Instead, there was significantly worse balance performance at T2 compared to T1 in the OAHL in the control condition with lower MoCA levels and the OA in the EF training condition at all MoCA levels (low, average, and high; see S2 for full statistics). To summarize, these interactions did not provide evidence of any unique training-related improvements to balance that were predicted by interactions among group-related factors (age and hearing status) and cognitive abilities.

#### Auditory 2-back performance

First, to address our primary objective of investigating the effects of EF training across Time and Groups on auditory-cognitive performance, an LMM was conducted with the auditory 2-back RTW (ms) as the dependent variable, for which higher values indicate worse performance. Results demonstrated a significant four-way Intervention*Time*Age*PTA interaction on 2-back RTW performance, *F*(1,418.43) = 4.700, *p* = .031, *η_p_ ^2^* = .01. Post-hoc tests revealed that for those in the EF training condition (but not the control condition) RTW at T2 was significantly better (faster) than at T1 for participants of average ages and lower PTAs (better hearing; *t* = 3.576, *p* < .001, see Fig 4). Further, for those in the EF training condition (but not the control condition), RTW at T2 was significantly better than at T1 for participants of older ages and lower PTAs (i.e., better hearing; *t* = 3.646, *p* < .001). Those in the control condition did not improve from T1 to T2; in fact, their performance generally worsened in terms of auditory 2-back performance (see S3 for full statistics). To summarize, participants in the EF training condition of average-to-older ages and lower levels of hearing loss demonstrated significant improvements in auditory 2-back RTW, with the control condition showing no improvements.

**Fig 4.**
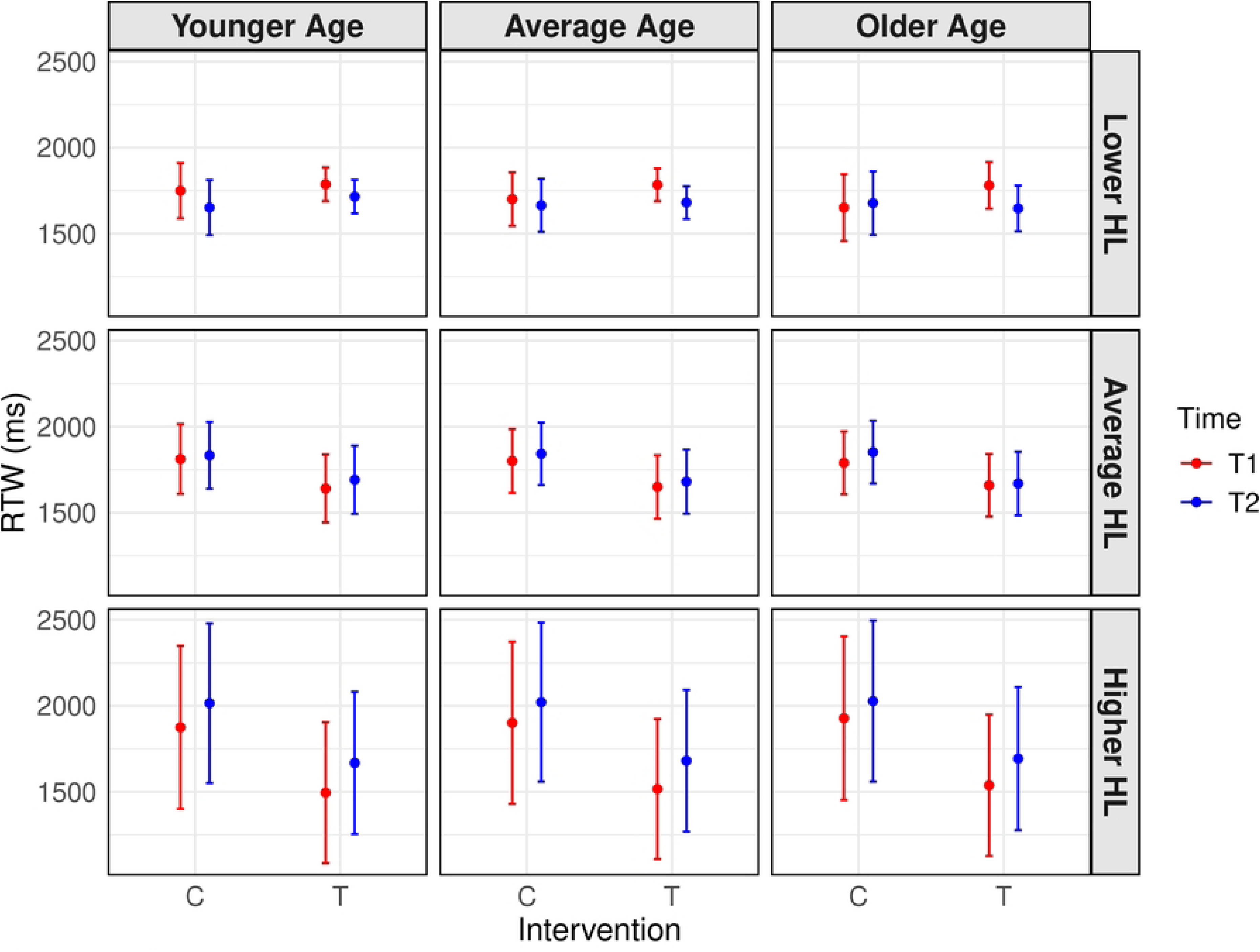
LMM of Intervention x Time x Age x PTA Interaction Effect on Auditory 2-back RTW.

**Fig 4**. Estimated marginal means of auditory 2-back reaction time weighted (RTW in ms, higher scores represent worse performance) from T1 to T2 for both the EF training (T) and control (C) conditions. Age and hearing loss (HL) tertiles are represented by the columns and rows respectively. All significant findings are stated in-text. Errors bars represent the 95% confidence intervals around model-estimated marginal means, which may extend beyond observed data ranges (i.e., max of 2000 ms) due to statistical estimation.

We also found a significant four-way Intervention*Time*Group (MA, OA, OAHL)*MoCA interaction on 2-back RTW performance, *F*(2,425.46) = 8.7397, *p*< .001, *η_p_ ^2^* = .04. Post-hoc tests demonstrated that for the EF training condition (but not the control condition) RTW was better at T2 than T1 for OAHL with lower MoCA scores (*t* = 2.145, *p* = 0.033). In contrast, OA at lower-average MoCA levels in the EF training condition had poorer RTW at T2 compared to T1 (see S3 for full statistics). The control condition did not improve from T1 to T2; in fact, they generally worsened in terms of auditory 2-back performance (see S3 for full statistics). To summarize, participants in the EF training condition with the worst hearing abilities (i.e., OAHL) and poorest cognitive abilities demonstrated significant improvements in auditory 2-back RTW, whereas the participants with normal hearing and lower-to-average cognition did not improve. Again, the control condition showed no improvements.

## Discussion

The current study evaluated whether EF training could improve cognitive and/or balance performance on an auditory-cognitive-while-balancing dual-task within a complex, realistic environment and whether any training-related improvements would differ as a function of hearing abilities and age. Results demonstrated that nearly all participants across the three age and hearing groups improved over the course of the training program on two of the three directly trained tasks (Stroop and visual *n*-back), demonstrating that the EF training program was generally successful for near transfer. The effects of this training also appeared to transfer to the primary auditory-cognitive and balance-related outcomes, but were influenced by age, hearing abilities, and cognitive abilities. In contrast, participants assigned to the control condition did not show any improvements in the primary outcomes from T1 to T2, and in some cases demonstrated significantly worsening performance. Of primary interest was whether or not EF training improved balance performance as a way of evaluating whether interventions to improve cognitive processing could improve mobility-related outcomes, particularly in older adults with hearing loss.

Results demonstrated that, for balance performance, there were significant training-related improvements for participants at older ages and with better hearing. However, for those with poor hearing, no training-related improvements were observed at any age. This suggests that once hearing loss is too severe, any training-related improvements in cognitive processing may not be sufficient to translate into better managing balance-related outcomes during realistic dual-tasks with auditory-cognitive demands. In contrast, for adults who are older, but who have better preserved hearing, EF training can indeed result in better balance-related performance outcomes when there are auditory-cognitive dual-task demands. An important consideration is that the PTA values in our study ranged from 3 to 60 dB HL (with one participant having a better-ear PTA of 71 dB HL). Based on standardized World Health Organization [92] classifications of hearing loss, where no impairment is defined as ≤25 dB HL, slight impairment as 26–40 dB HL, moderate impairment as 41–60 dB HL, and severe impairment as 61–80 dB HL, the participants with hearing loss in this study fall largely within the moderate impairment range, noting that for hearing aid users these PTAs reflect unaided thresholds, but they completed the experimental tasks while aided.

In terms of the training-related effects on auditory-cognitive task performance, we found significant improvements for participants of average-to-older ages and better hearing abilities. When considering how these factors interacted with cognitive abilities (MoCA), we observed improvements in participants with poorer hearing (i.e., OAHL) who also had the poorest cognition, whereas participants who had normal hearing (i.e., OA) and better cognition (lower-to-average) did not improve. This suggests that participants who had the most to gain (or the most room for growth) from EF training demonstrated improvements (i.e., OAHL with poorer cognition), whereas participants with perhaps less to gain (OA with normal hearing and better cognition) did not improve. Overall, these findings highlight the potential for EF training to support mobility and cognitive functioning in older adults but also suggest that more severe hearing loss may limit the effectiveness of such interventions on balance-related outcomes, emphasizing the potential importance of early intervention to support mobility.

When comparing the current study results to previous studies that have used the same EF training program [i.e., 70, 116] or a subcomponent of the program (i.e., visual discrimination task only in Bruce et al. [76] and Pothier et al. [71]), comparable cognitive and balance-related benefits are observed. Specifically, Downey et al. [70] examined the effects of EF training, aerobic exercise, or gross motor exercises (i.e., walking, coordination, balance exercises) on single- and dual-task performance (2-back and gait) in healthy older adults (hearing status unknown). They observed a large improvement in dual-task auditory-cognitive accuracy [70]. Similarly, in the current study, training-related improvements were also observed for auditory-cognitive task performance, specifically in participants with better hearing and average-to-older ages, and in OAHL with poorer cognition. Interestingly, Downey et al. [70] also found that participants with poorer cognition demonstrated more improvements in dual-task auditory-cognitive accuracy. The current study expands these findings by suggesting that hearing abilities can interact with cognitive abilities to differentially influence the effects of EF training on auditory-cognitive outcomes. In terms of balance and mobility-related outcomes, while Downey et al. [70] did not observe any training-related improvements in gait speed (m/s); in contrast, the current study observed training-related improvements in postural outcomes (reduced postural sway variability), specifically for participants of older ages and with better hearing. Since the Downey et al. [70] study recruited healthy participants with respect to cognitive and motor abilities, but did not examine hearing status, it is possible that differences in participants’ hearing statuses across studies related to differences in training-related benefits to mobility. Further the two studies differed in the types of balance/mobility tasks used, with standing balance evaluated in the current study and dynamic walking evaluated in Downey et al. [70]. Perhaps, during more complex dynamic assessments of mobility, training-related gains are less robust.

In Downey et al. [116] MA and OA completed the same 12-week at-home EF training program used in the current study. Effects on single- and dual-task performance on an auditory-cognitive task (2-back task) and a mobility task (treadmill walking) and on patterns of brain activity using functional near-infrared spectroscopy were examined. The results showed that both the MA and OA groups with greater baseline cognitive processing (i.e., better MoCA scores) demonstrated improvements after EF training; specifically, reductions in dual-task costs were observed on both tasks [116]. In the current study, we demonstrated significant improvements on most of the EF training tasks (i.e., *n*-back, Stroop), with the results indicating training-related benefits to cognition (i.e., those with worse cognition and hearing showed training-related benefits).

Pothier et al. [71] examined the effects of exercise and/or cognitive training on spontaneous walking speed in healthy older adults (hearing status unknown). Overall, they found faster spontaneous walking speed in participants who received at least one active intervention compared to the control. Similarly, in the current study we also found training-related increases in postural stability in healthy older adults, but specifically those with better hearing. Bruce et al. [76] evaluated the effects of exercise training and cognitive training implemented simultaneously or sequentially in older adults with and without hearing loss. In their study the primary outcomes were performance on four different balance/mobility tasks (sit-to-stand task, and computerized dynamic posturography of double support, visual sway-referenced, and single-support) when performed alone and simultaneously with a 1-back task. They demonstrated that both older adults with and without hearing loss improved in sit-to-stand performance under dual-task conditions, but not on the other balance-related outcomes [76]. They also demonstrated training-related improvements in single- and dual-task cognitive performance (especially when task demands were low). These results are partially consistent with the current study given that both studies observed improvements in auditory-cognitive performance following training. However, whereas Bruce et al. [76] did not observe any improvements in postural outcomes (double support, visual sway-referenced, and single-support), the current study observed improvements in postural stability (reduced APSD), primarily for older adults with better hearing. There are a few differences between the Bruce et al. [76] study and the current study that are important to consider. Specifically, the Bruce et al. [76] recruited OANH (*M_age_* = 67 year) and OAHL (*M_age_* = 70 years) who were, on average, younger than participants in the current study (*M_age_* of OANH = 71 years, *M_age_* of OAHL = 77 years). Further, Bruce et al. [76] included an OAHL group with mild hearing loss (between 26-40 dB HL), whereas the current study included OAHL in both the mild and moderate hearing loss ranges (i.e., 3-71dB HL) with majority of the participants having moderate hearing loss when unaided and all of whom wore hearing aids. Although, both studies recruited those with normal cognition, the MoCA cut-offs were different with the Bruce et al. [76] cut-off being 26 or more out of 30 [97] and the current study cut-off being 23 or more out of 30 [97–98]. Further, Bruce et al. [76] implemented sit-to-stand and dynamic posturography tasks, while the current study implemented a static standing balance task. These differences suggest that EF training-related benefits to postural outcomes may differ as a function of age, hearing loss, cognitive abilities and difficulty of balance-related task demands; specifically, participants who are younger, with milder hearing loss, and better cognition may not benefit as much from EF training, particularly during more dynamic postural tasks.

### Cognitive aging frameworks to understand dual-task auditory-cognitive and balance performance

Several conceptual frameworks of cognitive aging have been proposed that can be referenced to better understand the factors likely to influence the extent to which cognitive training-related effects are observed. One framework largely includes two key mechanisms, compensation and magnification [118]. *Compensation* includes the following principles: 1) individuals with lower baseline performance can benefit the most from cognitive training through compensation and 2) there is a negative correlation between baseline performance/cognitive abilities and cognitive training gains [118]. There is some evidence supporting compensation in the current study for both balance-related outcomes and auditory-cognitive related outcomes. For example, training-related improvements to balance were observed only for participants who were in the older age range and who therefore might have more to gain from training compared to participants who were younger. Similarly, training-related improvements to auditory-cognitive task performance were mostly observed for older adults with hearing loss who had poorer cognition who may have had more to gain from cognitive training than those with normal hearing and better cognition. In contrast, *magnification* refers to a pattern in which individuals with higher cognitive processing gain more from training due to an increased ability to acquire and refine cognitive strategies and includes the following principles: 1) individuals with higher baseline performance derive greater benefits from training than those with lower baseline performance and 2) there is a positive correlation between baseline performance/cognitive abilities and cognitive training gains [118]. There is some evidence supporting magnification in the current study as the training-related improvements to both primary outcomes (RTW, APSD) were found in the individuals with better hearing, but not in those with poorer hearing.

For the balance outcomes, older adults with better hearing benefited the most from training compared to poorer hearing at any age. For auditory-cognitive task performance, average to older adults with better hearing benefited from training, while those with the poorest cognition and poorer hearing benefited the most from training. Overall, this pattern of results suggests that there may be some processing limit for hearing that was reached, such that beyond a certain degree of hearing loss, cognitive training may no longer be effective, or more cognitive training may be required to improve balance during realistic dual-tasks with auditory-cognitive demands. However, as demands increase, eventually a resource-limited ceiling is reached, leading to an inability to compensate, leading to associated behavioural performance declines and perhaps associated declines in training-related benefits. In the current study, it is possible that once an upper threshold for hearing loss is reached, EF training may no longer be sufficient to compensate for the high demands required to perform an auditory-cognitive task while maintaining standing balance.

### Limitations and future directions

An important limitation of the current study is the nature of the participant samples. We recruited generally healthy, high functioning adults with no major health conditions or other comorbidities that are common with older age and other health conditions that increase risk for cognitive decline and hearing loss (e.g., diabetes). For some measures, including the MoCA, we had a very small range of abilities that were limited to the better end of the ability spectrum. While this strategy of only recruiting physically and cognitively healthy participants was intentional to limit potential confounds in the interpretation of the data, it also limits the generalizability of these findings to the broader older adult population, which is highly heterogeneous. Consequently, the current results likely provide a conservative estimate and might not adequately reflect the potential magnitude of EF training benefits on cognitive-mobility performance in the general population or in real-world settings. Future research would benefit from broadening eligibility criteria to include adults with a wider range of sensory, cognitive, and mobility abilities to better evaluate the extent to which these abilities predict training-related outcomes.

Efforts were made to achieve a balanced representation of sex across study groups; however, dividing participants by intervention conditions (EF training vs. control) yielded subgroup sizes too small to permit meaningful analyses of sex-related differences. Nonetheless, sex and gender are important factors to consider, as both may influence the efficacy of EF training on cognitive and sensorimotor performance. Moreover, hearing loss tends to be more prevalent in men than in women, a disparity attributed to both biological and environmental influences [119–121]. For example, estrogen is thought to exert a protective effect on auditory function in premenopausal women (i.e., a sex-related factor), while greater exposure to occupational noise in male-dominated industries (e.g., construction, manufacturing) may contribute to elevated risk among men (i.e., a gender-related factor; 119-121). Future studies should recruit larger, stratified samples to examine how these sex and gender differences may interact with auditory, cognitive, and balance outcomes [122–124].

Another potential limitation of the current study is the absence of long-term follow-up assessments to evaluate the sustainability of the observed EF training effects on auditory-cognitive and balance performance. While short-term post-training improvements were observed in both auditory-cognitive and postural measures in the current study, it remains unclear whether these gains could be sustained over time. Without longitudinal data, we cannot determine the extent to which the observed benefits reflect temporary performance boosts versus lasting functional improvements. Of note are two studies [125–126] that examined the long-term effects and durability of EF training on cognitive abilities and everyday functions in independently living older adults. Both studies trained memory, reasoning, and speed of processing separately and demonstrated training resulted in fewer declines to independent activities of daily living compared to the control condition [125], fewer declines to independent activities of daily living after reasoning training specifically [126] these gains were sustained for at least 5 years [126]. However, these longitudinal studies did not consider the effects of hearing loss on observed benefits to independent activities of daily living. Future studies should incorporate follow-up assessments to evaluate the durability of cognitive training effects on balance and cognitive outcomes over time and in individuals with hearing loss. Important future considerations to determine the efficacy of at-home executive function training can include the length of time spent training, the time and intensity of training (e.g. time per day/days per week), and types of cognitive domains trained. Moreover, longitudinal studies are also required to determine whether any training-related improvements in balance and/or auditory-cognition also result in real world outcomes such as reduced fall risk or improved mobility.

### Conclusions

The current study expanded on the existing literature examining the fundamental interactions among hearing, cognition, and mobility across the middle-aged and older adult lifespan. Specifically, we assessed whether EF training benefits would be observed on balance and auditory-cognitive task performance and whether these training-related effects would be predicted by age, hearing status, and standardized sensory, cognitive, and mobility outcomes. Over the course of the 12-week EF training, participants improved on measures of working memory and inhibitory control and task-switching abilities but not divided attention. For the balance outcomes, performance for older adults with better hearing improved after training; those with poorer hearing (at any age) did not improve. For auditory-cognitive task performance, adults of average to older ages with better hearing improved after training; however, when cognitive abilities were considered, those with the poorest cognition who also had the poorest hearing benefited the most from training. These findings contribute to what is known about the interactions among hearing loss, cognition, and mobility during complex, realistic activities and how EF training can potentially improve cognitive processing to support hearing and mobility across the adult lifespan.

## Data Availability

Data cannot be shared publicly because of the KITE-Toronto Rehabilitation Institute-University Health Network's confidentiality agreement for data sharing policies. Data are available from the KITE-Toronto Rehabilitation Institute-University Health Network's Research Ethics Board for researchers who meet the criteria for access to confidential data.

## Acknowledgments

We would like to thank Colin Stoddart, Roger Montgomery, Dr. Bruce Haycock and Susan Gorski for their technical assistance, Joanne Nuque, Kristen Arnold, and Sumayah Herzi with assistance in collecting and transcribing data and Lauryn Gittens for support in transcribing data and making Table 1.

Photo Credit: Tim Fraser.

## Supporting Information

**S2 Fig. LMM of Intervention x Time x Group x MoCA Interaction Effect on COP APSD**

**S2 Fig.** Estimated marginal means of Centre of Pressure Path Length Anterior-Posterior Standard Deviation (COP APSD in mm; higher scores represent worse performance) from T1 to T2 for those in both the executive function cognitive training (T) and control (C) conditions. Group and Montreal Cognitive Assessment (MoCA) tertiles are represented by the columns and rows respectively. Errors bars represent the 95% confidence intervals.

**S3 Fig. LMM of Intervention x Time x Group x MoCA Interaction Effect on Auditory 2-back RTW**

**S3 Fig.** Estimated marginal means of auditory 2-back Reaction Time Weighted (RTW in ms, higher scores represent worse performance) from T1 to T2 for both the Executive Function training (T) and control (C) conditions. Group and Montreal Cognitive Assessment (MoCA) tertiles are represented by the columns and rows, respectively. Errors bars represent the 95% confidence intervals around model-estimated marginal means, which may extend beyond observed data ranges (i.e., max of 2000 ms) due to statistical estimation.

